# Reliance on vision for walking balance is related to somatosensory deficits in individuals with cerebral palsy

**DOI:** 10.1101/2024.02.07.24302467

**Authors:** Ashwini Sansare, Hendrik Reimann, Barry Bodt, Maelyn Arcodia, Khushboo Verma, John Jeka, Samuel C.K. Lee

**Author notes:** **Correspondence:** Samuel C.K. Lee.

## Abstract

**Aim:** To investigate the relationship between somatosensory deficits, specifically ankle and hip joint position sense, two-point discrimination, and vibration on the (1) responses to visual perturbations during walking and (2) response improvements to visual perturbations while receiving a sensory-centric treatment, i.e., stochastic resonance (SR) stimulation, in individuals with and without cerebral palsy (CP).

**Methods:** Twenty-eight individuals (14 CP, 14 age-and sex-matched controls) walked in a virtual reality cave while receiving visual perturbations. We applied SR to the ankle and hip joints. Data analysis consisted of bivariate correlations, and multiple regression analysis (MRA) using all four sensory tests as predictors with the responses to visual perturbation and the improvements in the responses when SR is applied as outcomes.

**Results:** We found significant and strong correlations between performance on sensory tests and the responses to visual perturbations, and improvements in the responses with SR. Only one predictor could be entered into the MRA, indicating that performance on any of the sensory tests could predict the responses to visual perturbation and the improvements with SR.

**Interpretation:** Individuals with sensory deficits are more responsive to sensory-centric interventions. This study is an initial step in identifying potential “responders” to sensory therapies in individuals with CP.

## 1. INTRODUCTION

Postural instability is the most significant contributor to the primary impairments in cerebral palsy (CP) (Jeffries et al., 2016), with about 35% reporting daily falls and 30% reporting weekly or monthly falls(Boyer & Patterson, 2018). Falls can occur because of deficits in either motor or sensory systems. Most balance assessments in individuals with CP typically focus only on the motor aspect, with little to no consideration of how sensory deficits might impact balance. However, there is increasing evidence that links sensory deficits, both central and peripheral, to balance control and walking performance in CP. Mobility impairments in CP, including slower walking speed and shorter steps, have been associated with abnormal somatosensory cortical activity (Kurz et al., 2015). Additionally, poor performance on clinical sensory tests of lower extremity, such as aberrant two-point discrimination, light touch, hip and ankle joint position sense have been found to have moderate to strong relationships with reduced balance parameters such as increased postural sway, poor BESTest scores and reduced walking performance, indicated by slower gait speed, shorter step length and shorter 6 Minute Walk Test distance (Damiano et al., 2013; Zarkou et al., 2020). In summary, these findings suggest a strong link between somatosensory deficits and postural control in CP.

Individuals with CP rely on visual information over other sensory modalities for regulating postural sway. With walking, our previous work has shown that visual perturbations led to a magnified and delayed body sway in response to the perturbations in the CP group compared to their age-and sex-matched peers. Other studies have shown similarly increased postural sway during standing in children with CP(Barela et al., 2011; Yu et al., 2018) and during walking in older adults (Franz et al., 2015) when their visual input was manipulated. The central hypothesis to explain the increased sway in these studies is that individuals with CP compensate for somatosensory deficits, particularly in proprioception, by relying on vision for balance, rather than information from the impaired somatosensory system. Hence, these individuals are more likely to be affected by changes in visual information, leading to the observed larger body sway response to visual perturbations. However, there is currently no direct evidence for this hypothesis as none of the prior work has directly investigated the link between response to visual perturbations and somatosensory deficits.

The hypothesis that visual reliance is a compensation for somatosensory deficits also implies that those with greater somatosensory deficits will show greater improvement with interventions that improve somatosensory processing for balance control. In other words, individuals with degraded somatosensation should respond better to interventions that reduce reliance on vision One intervention that improves somatosensation is stochastic resonance (SR) stimulation. SR involves applying random, sub-sensory noise to sensory receptors, which in turn causes small changes in the transmembrane potentials and makes the receptors more likely to fire an action potential(Cordo et al., 1996; Moss et al., 2004; Ribot-Ciscar et al., 2013). Our recent work investigated how the responses to visual perturbations during walking would change in individuals with CP when applying SR to improve proprioceptive input(Sansare et al., 2022). Our results showed that the CP group had reduced whole-body sway when receiving SR stimulation. These results, along with other studies(Gravelle et al., 2002; Priplata et al., 2006; Ross et al., 2013; Ross & Guskiewicz, 2006) that have used SR or similar means of improving sensory input to improve balance, indicate that augmenting somatosensory input helps individuals with CP reduce their reliance on vision, thus showing a reduced response to visual perturbations. More importantly, SR showed greater improvements in standing body sway in young healthy individuals with greater baseline sway. In another study on the effects of SR on gait variability in older adults, SR reduced gait variability in participants who already had a variable gait, but increased variability in participants with the least baseline variability(Stephen et al., 2012). Thus, improvements in balance control after receiving a sensory-centric intervention such as SR seem to be related to baseline functional abilities. However, it is not known whether there is a relationship between the level of somatosensory deficit and the corresponding response to sensory augmentation.

Here we performed a secondary analysis of our prior studies (Sansare et al., 2022, 2023; Sansare Ashwini et al., n.d.) in individuals with CP and age-and sex-matched typically developing peers (TD), with two objectives : (1) to investigate the relationship across subjects between their response to visual perturbations during walking and their somatosensory deficits, specifically ankle and hip joint position sense, two point discrimination, and vibration, and (2) to investigate the relationship between the individual improvement in the response to visual perturbations from SR stimulation and their somatosensory deficits. We hypothesized (1) that individuals with greater responses to visual perturbations would have greater somatosensory deficits, and that (2) individuals with greater improvement in the response to visual perturbations would have greater somatosensory deficits.

## 2. METHODS

The study is a secondary analysis of the data collected in a previously reported study, wherein day one investigated the baseline differences in the response to visual perturbations between individuals with and without CP (Sansare et al., 2022), and day two of the study (registered at clinicaltrials.gov under NCT05233748), investigated the immediate improvement in the response to visual perturbations while receiving augmented proprioception through SR stimulation in individuals with and without CP(Sansare et al., 2023).

### 2.1 Participants

Fourteen ambulatory individuals with spastic or hemiplegic CP, with Gross Motor Function Classification System (GMFCS) levels I–II were recruited through the CP Clinic at Nemours Childrens Hospital, Delaware, and community flyers. Fourteen age-matched (±6 months) and sex-matched typically developing (TD) individuals were recruited through flyers, local advertisements, and social media. The Institutional Review Board at the University of Delaware approved the study protocol. Informed parental consent and child assent were obtained. The inclusion and exclusion criteria are listed in Table 1.

**Table 1.**
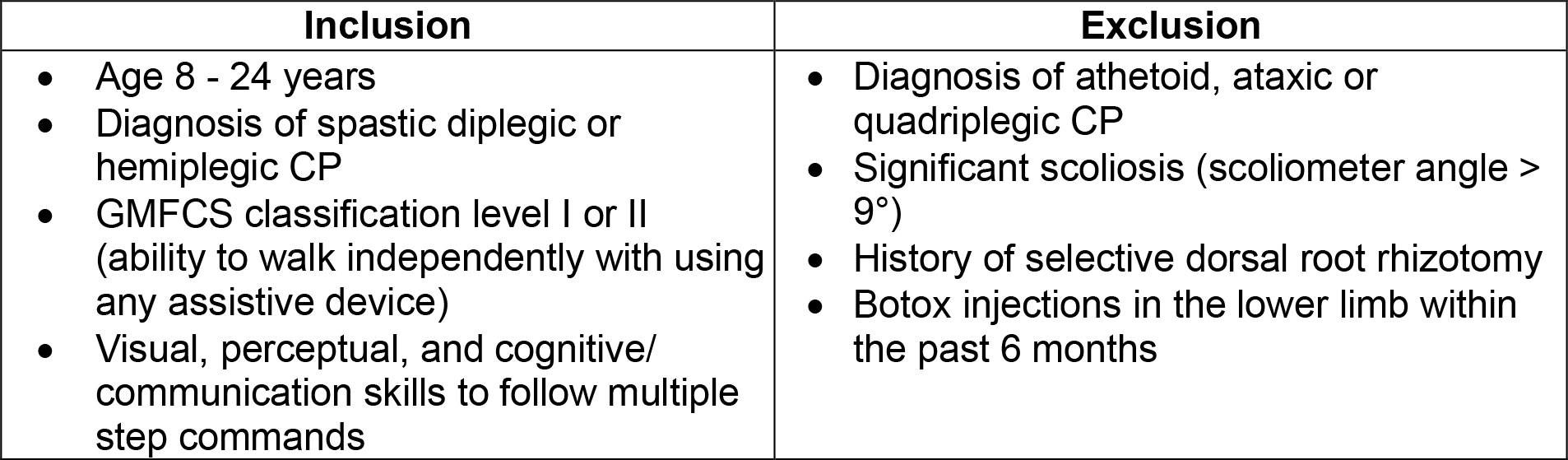
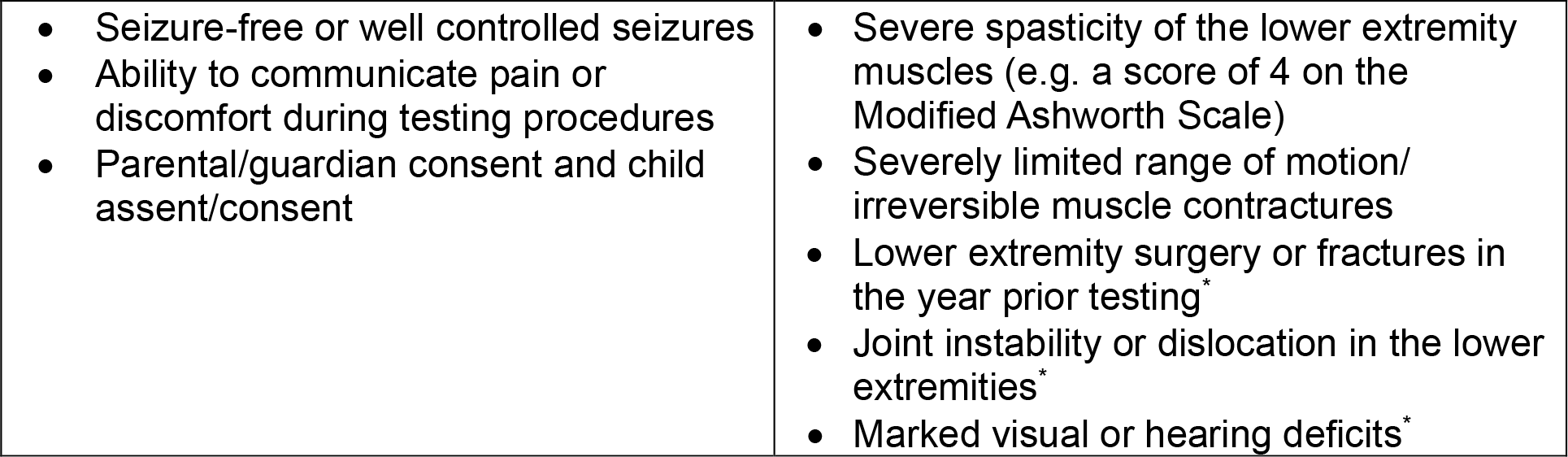
Inclusion and exclusion criteria. The asterisk denotes criteria applicable to TD group.

| Inclusion | Exclusion |
| --- | --- |
| • Age 8 - 24 years<br>• Diagnosis of spastic diplegic or hemiplegic CP<br>• GMFCS classification level I or II (ability to walk independently with using any assistive device)<br>• Visual, perceptual, and cognitive/communication skills to follow multiple step commands | • Diagnosis of athetoid, ataxic or quadriplegic CP<br>• Significant scoliosis (scoliometer angle $> 9^\circ$)<br>• History of selective dorsal root rhizotomy<br>• Botox injections in the lower limb within the past 6 months |
| • Seizure-free or well controlled seizures<br>• Ability to communicate pain or discomfort during testing procedures<br>• Parental/guardian consent and child assent/consent | • Severe spasticity of the lower extremity muscles (e.g. a score of 4 on the Modified Ashworth Scale)<br>• Severely limited range of motion/irreversible muscle contractures<br>• Lower extremity surgery or fractures in the year prior testing*<br>• Joint instability or dislocation in the lower extremities*<br>• Marked visual or hearing deficits* |

### 2.2 Measures

#### 2.2.1 Somatosensory tests

Joint position sense: The joint position sense was evaluated at the ankle (dorsiflexion and plantarflexion), and hip joints (flexion). A physical therapist moved the distal segment of the joint being tested into a target position, held it in that position for three seconds before moving it back to neutral position. The participant was then asked to actively reproduce the same motion as accurately as possible on the same limb. The magnitude of error between the target position by the therapist and the reproduction by the participant was recorded for each joint (average over three trials)(Wingert et al., 2009).

Two-point discrimination: An aesthesiometer (Baseliner, White Plains, New York, NY, USA) at the first metatarsal head on the plantar aspect of each foot was used to assess two-point discrimination. Scores consisted of the minimum distance (cm) between two stimulus points, which were correctly identified as two distinct points on at least two out of three trials(Bell-Krotoski et al., 1993).

Cutaneous Vibration: A 128 Hz tuning fork (Rydel–Seiffer graduated tuning fork, Martin, Tuttlingen, Germany) at the first metatarsal head on the plantar aspect of each foot was used to assess cutaneous vibration. The participant was asked to let the therapist know once vibration was perceived to dissipate completely and time was recorded as the average duration that the participant perceived over three trials. The shorter the time for which the vibration was perceived, the worse the sensory impairment(Bell-Krotoski et al., 1993; Temlett, 2009).

#### 2.2.2 Response to visual perturbations

Participants walked in a virtual reality environment on a self-paced treadmill. The visual perturbations were induced by intermittently tilting the virtual scene randomly every 10-12 steps to induce a sensation of a sideways fall. After completing two 2-minute practice trials to acclimatize to the treadmill and the virtual environment, each participant completed ten 2-minute walking trials with visual perturbations. We measured the response to visual perturbations as area-under-the-curve of the CoM, where higher values imply a magnified response to the visual perturbations and, in turn, a greater reliance on vision for walking balance. For brevity, we will refer to this measure simply as “CoM excursion”. For further details please refer to Sansare et al(Sansare et al., 2022).

#### 2.2.3 Improvement in response to visual perturbations

To investigate the effect of SR on the response to the visual perturbations we applied SR stimulation at the optimal intensity for each individual (SRopt) at the hips, lower legs and ankles, as well as a control condition without stimulation (noSR) while walking. Participants completed a total of six walking trials, with three trials of two minutes each under the SRopt and noSR conditions. They received visual perturbations in the same manner as described above. For further details of the SR stimulation protocol please see the primary analysis (Sansare Ashwini et al., n.d.). Our primary outcome measure to quantify the improvement in the response to visual perturbations was the difference in the CoM excursion between the SR and the noSR trials, where a positive value indicates that the CoM excursion reduced with SR, implying an improvement in the response to visual perturbations.

### 2.3 Data Analysis

We tested whether the four sensory tests, (1) ankle joint position sense, (2) hip joint position sense, (3) vibration, and (4) two point discrimination, can predict (a) the response to visual perturbation and (b) improvement with SR stimulation each with their performance on each somatosensory test. To determine the relationships between the four somatosensory predictors and the two outcomes, we performed bivariate correlations between all predictor-outcome pairs. Further, we conducted multiple regression analysis (MRA) using all four somatosensory predictors simultaneously. For model selection and assumptions for MRA, please refer to Supplementary File 1.

For both bivariate correlations and MRA, we analyzed the group (CP,TD), and the affected side (More Affected, Less Affected) separately, resulting in four separate cases (CP More Affected, TD More Affected, CP Less Affected and TD Less Affected). All analyses were conducted in JMP, (Version 17.0.0, SAS Institute Inc., Cary, NC). We determined the More affected side as the one with hemiplegia in individuals with hemiplegic CP, the one self-identified as the more affected side in individuals with diplegic CP, and the non-dominant side for individuals with TD. The dominant side in the TD group was self-determined by the participants as their preferred lower limb of use during daily activities.

## 3. RESULTS

### 3.1 Results from bivariate correlations

Descriptive statistics for performance on all the sensory tests for the More and Less Affected side for CP and TD are given in Table 2 below.

**Table 2.** Mean and standard deviation (SD) for CP and TD groups on the More Affected and Less Affected Side for the four outcome measures.

| Outcome Measure | More Affected Side |  |  |  | Less Affected Side |  |  |  |
| --- | --- | --- | --- | --- | --- | --- | --- | --- |
|  | CP |  | TD |  | CP |  | TD |  |
|  | Mean | SD | Mean | SD | Mean | SD | Mean | SD |
| Ankle Joint Position Sense (degrees) | 6.547 | 3.213 | 2.286 | 1.820 | 4.143 | 1.906 | 1.809 | 1.224 |
| Hip Joint Position Sense (degrees) | 3.451 | 1.369 | 1.832 | 1.806 | 2.809 | 1.389 | 1.404 | 1.178 |
| Two point discrimination (meters) | 1.45 | 0.277 | 0.857 | 0.224 | 1.157 | 0.122 | 0.771 | 0.270 |
| Vibration (seconds) | 8.822 | 2.407 | 12.130 | 2.849 | 9.295 | 2.966 | 11.960 | 2.631 |

#### 3.1.1 Response to visual perturbations

Figures 1 and 2 show bivariate correlations between the response to visual perturbations and the four sensory tests on the More and Less affected side, respectively, while Table 3 depicts the p values and correlation coefficients (r).

**Table 3.**
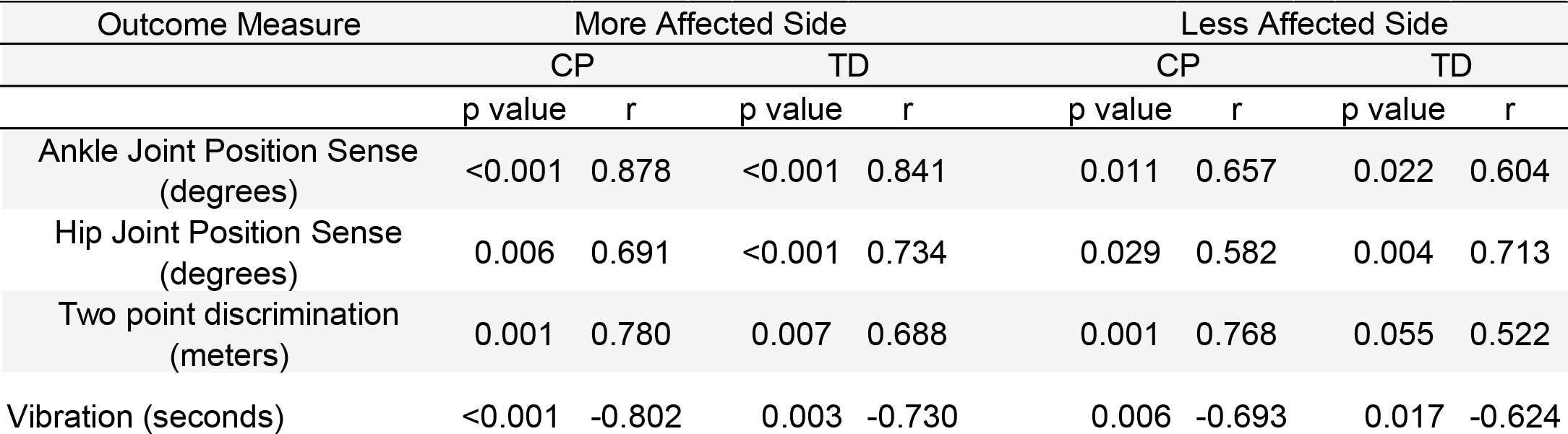
P values and R for CP and TD groups on the More Affected and Less Affected Side for bivariate correlations between response to visual perturbations and each of the four outcome measures.

**Figure1.**
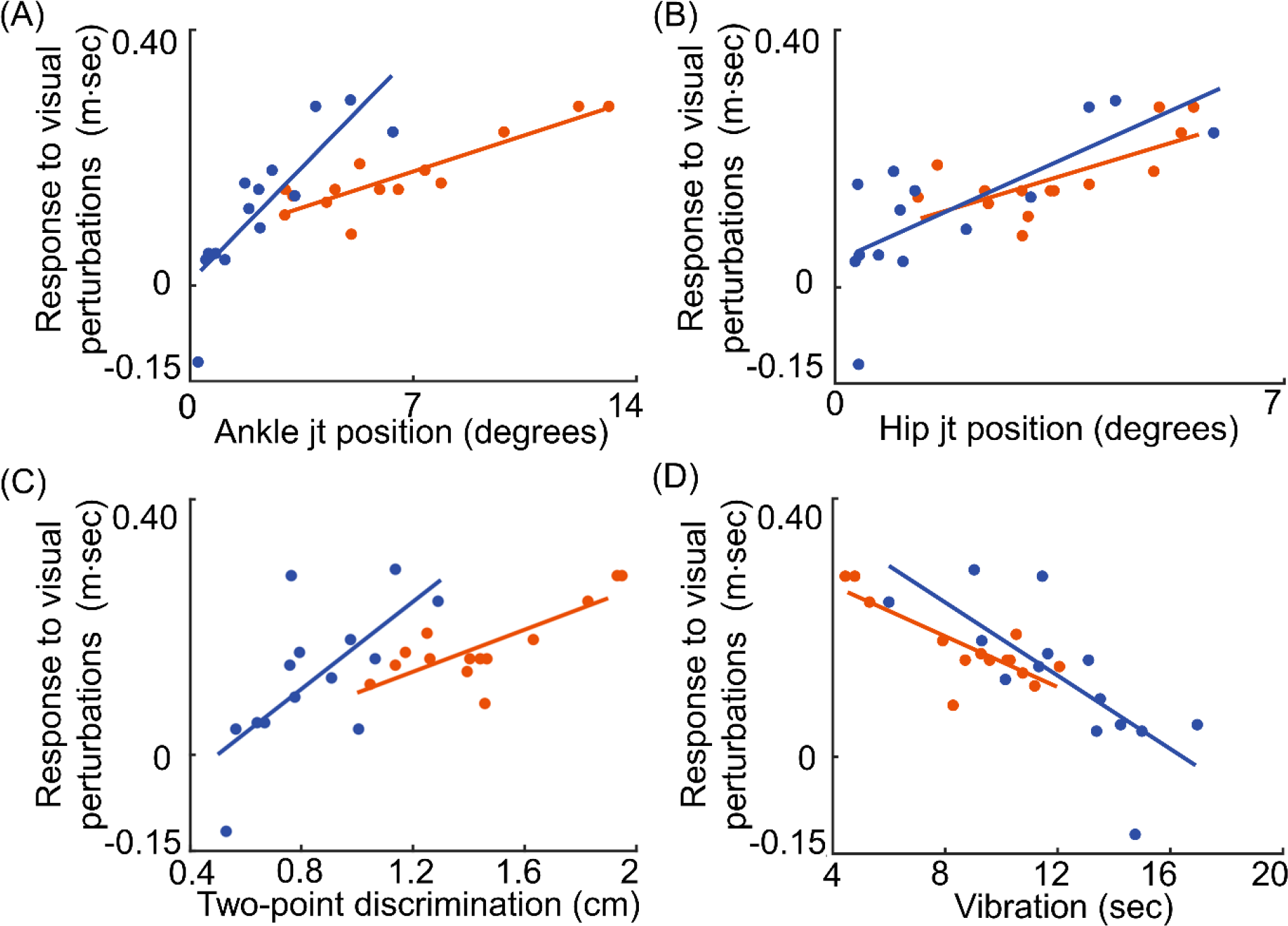
Bivariate correlations between the four somatosensory predictors (horizontal axes) and the CoM excursion in response to the visual perturbations (vertical axes) from the More affected side. The predictors are ankle joint position sense (Panel A), hip joint position sense (Panel B), two point discrimination sense (Panel C), and vibration sense (Panel D).CP is orange and TD blue.

**Figure 2.**
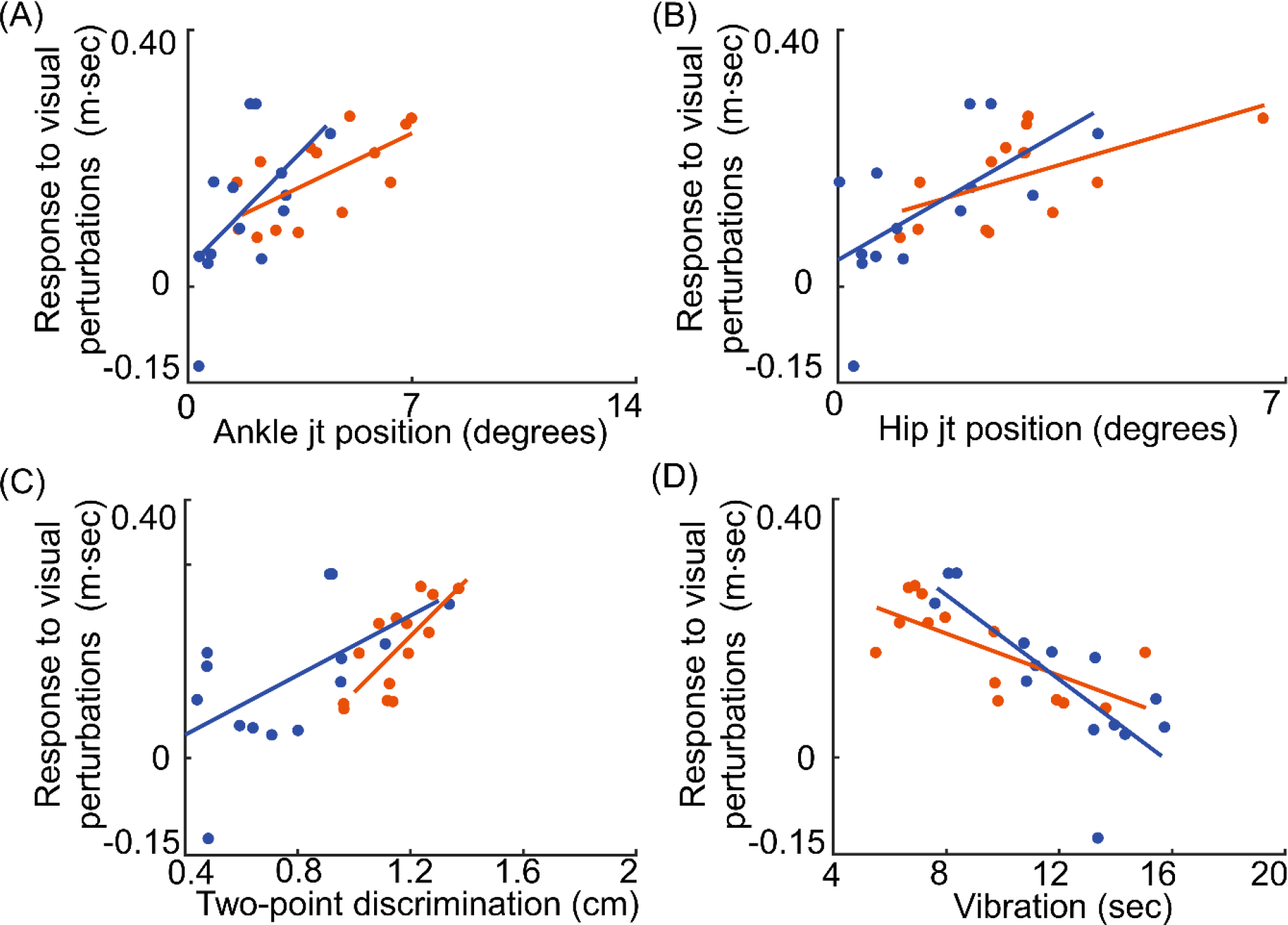
Bivariate correlations between the four somatosensory predictors (horizontal axes) and the CoM excursion in response to the visual perturbations (vertical axes) from the Less affected side. The predictors are ankle joint position sense (Panel A), hip joint position sense (Panel B), two point discrimination sense (Panel C), and vibration sense (Panel D). CP group is in orange and TD group is in blue.

Performance on all four sensory tests showed statistically significant pairwise correlations with the response to visual perturbations for both groups (CP and TD) and sides with the exception of two-point discrimination for the Less Affected side for TD group (p = 0.055,Table 2). Out of the four somatosensory tests, ankle joint position sense showed the strongest correlation (R= 0.878, p < 0.001) with the response to visual perturbations.

#### 3.1.2 Improvement in response to visual perturbations

Figures 3 and 4 show bivariate correlations between the response to visual perturbations and the four sensory tests on the More and Less Affected Side, respectively, while Table 4 depicts the p values and correlation coefficients (r).

**Table 4.** P values and R for CP and TD groups on the More Affected and Less Affected Side for bivariate correlations between improvement in response to visual perturbations and each of the four outcome measures.

| Outcome Measure | More Affected Side |  |  |  | Less Affected Side |  |  |  |
| --- | --- | --- | --- | --- | --- | --- | --- | --- |
|  | CP |  | TD |  | CP |  | TD |  |
|  | p value | r | p value | r | p value | r | p value | r |
| Ankle Joint Position Sense (degrees) | 0.006 | 0.692 | 0.284 | 0.308 | 0.128 | 0.427 | 0.062 | 0.511 |
| Hip Joint Position Sense (degrees) | 0.002 | 0.754 | 0.214 | 0.354 | 0.006 | 0.693 | 0.311 | 0.292 |
| Two point discrimination (meters) | 0.001 | 0.773 | 0.029 | 0.581 | 0.001 | 0.780 | 0.056 | 0.520 |
| Vibration (seconds) | <0.001 | -0.820 | 0.401 | -0.244 | 0.026 | -0.592 | 0.117 | -0.439 |

**Figure 3.**
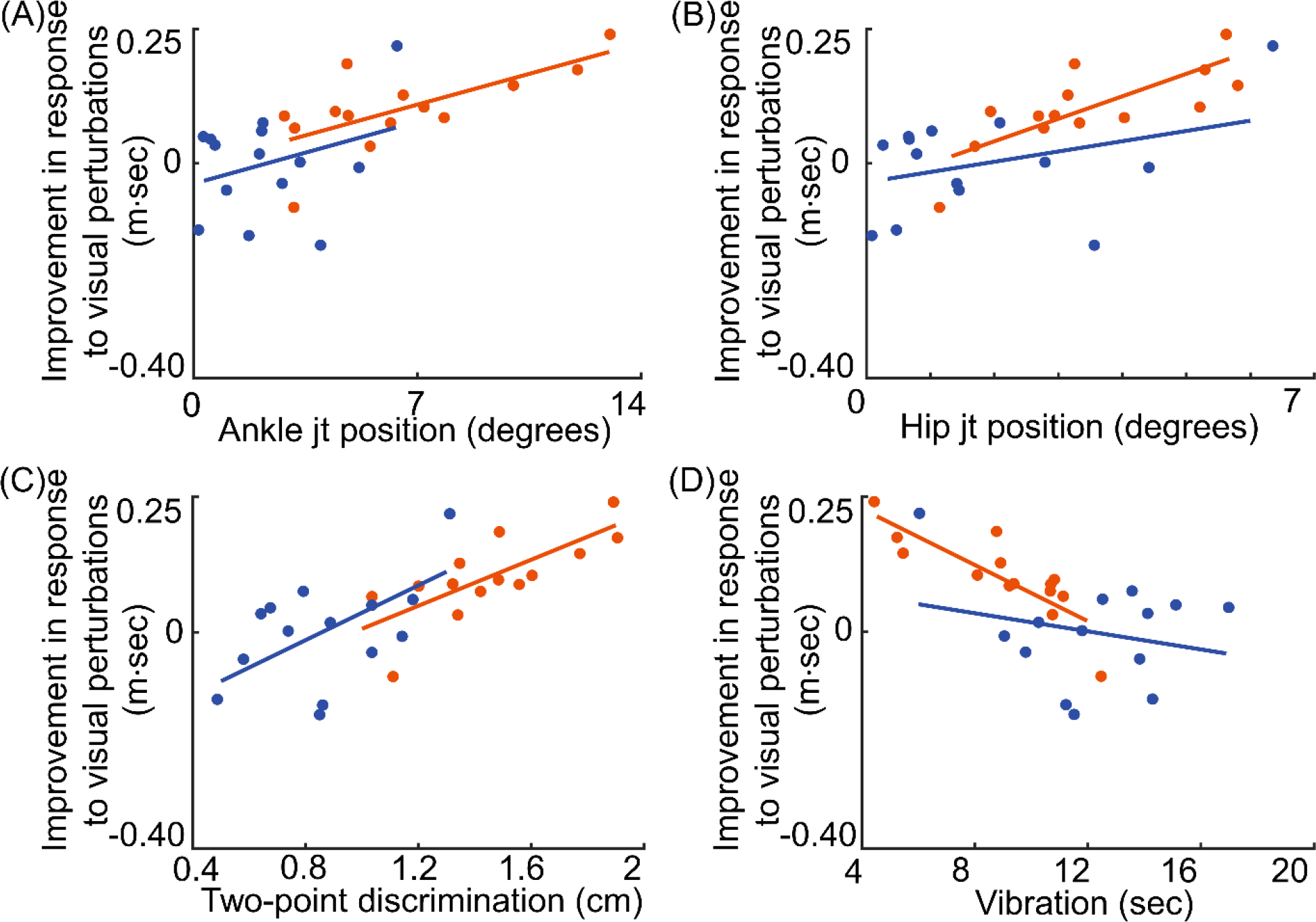
Bivariate correlations between the four somatosensory predictors (horizontal axes) and the improvement in response to the visual perturbations (vertical axes) from the More affected side. The predictors are ankle joint position sense (Panel A), hip joint position sense (Panel B), two point discrimination sense (Panel C), and vibration sense (Panel D). CP group is in orange and TD group is in blue.

**Figure 4.**
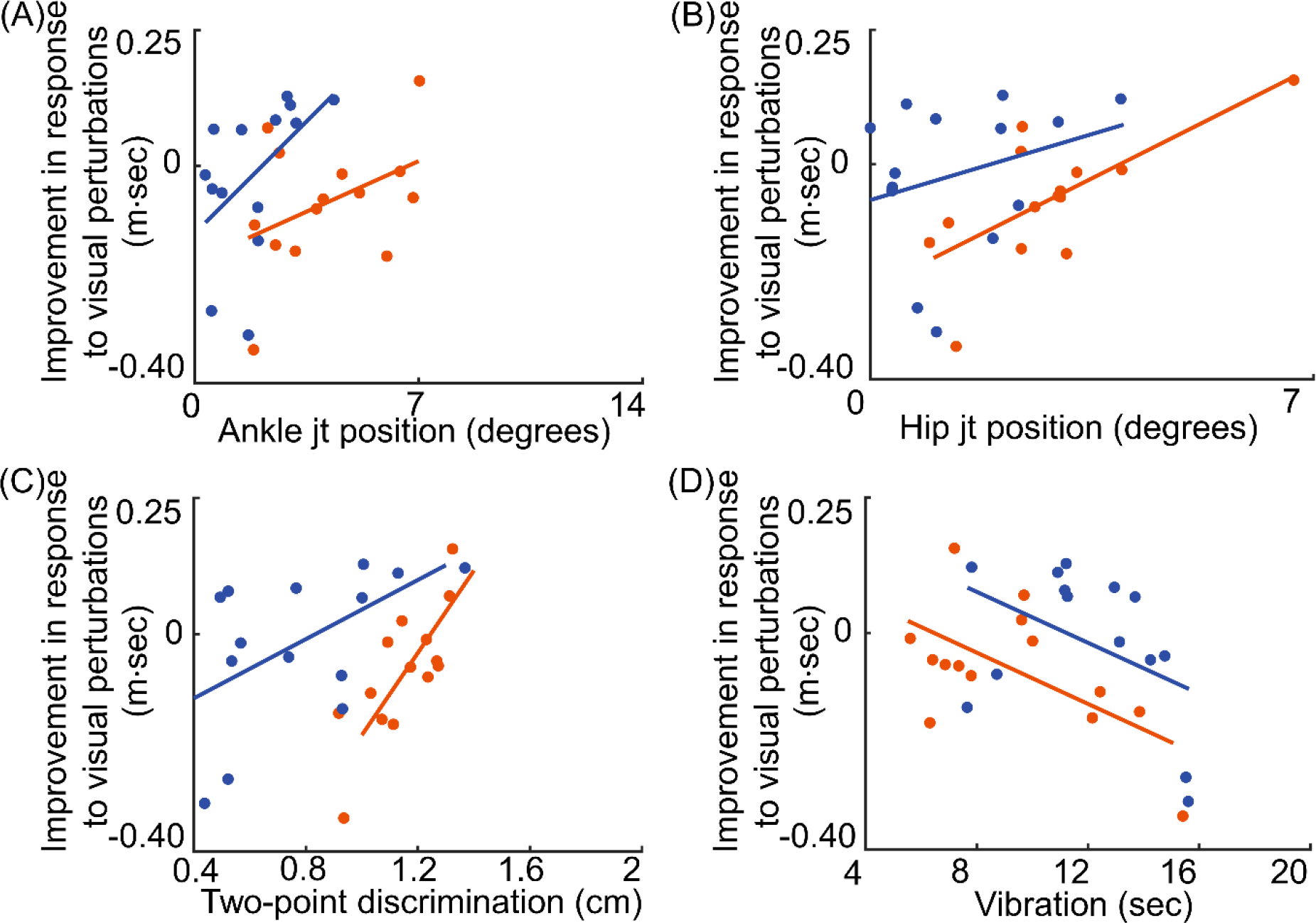
Bivariate correlations between the four somatosensory predictors (horizontal axes) and the improvement in response to the visual perturbations (vertical axes) from the Less affected side. The predictors are ankle joint position sense (Panel A), hip joint position sense (Panel B), two point discrimination sense (Panel C), and vibration sense (Panel D). CP group is in orange and TD group is in blue.

Performance on all four sensory tests showed a statistically significant correlation with improvement in the response to visual perturbations during SR stimulation. Out of the four somatosensory tests, vibration showed the strongest correlation (r= 0.820, p < 0.001) (Table 4) with the improvement in response to visual perturbations.

### 3.2 Results from multiple regression analysis

The goal of performing an MRA over and above the bivariate correlations was to account for all of the predictors in a single model, thus yielding an understanding of the associations between the predictors as a whole and the associations between the various predictors themselves. However, multicollinearity i.e., the correlation among the somatosensory predictors themselves was sufficiently high that including more than one predictor in any model did not substantially improve the model fit. Thus, with models effectively reduced to one predictor, significant models are supported by any of the four predictors for the response to visual perturbation regardless of More/Less Affected Side and CP/TD group, with the single exception that 2 pt discrimination fails to achieve significance for less affected side among TD. For the improvement in the response with SR, a similar claim can be made for CP, with the exception of ankle joint position sense for the Less affected side. Additional details are included in Supplementary File 1.

## 4. DISCUSSION

This study examined the relationship between the performance on somatosensory tests and (a) the responses to visual perturbations, and (b) the improvement in the response to visual perturbations during SR stimulation, an intervention designed to improve somatosensory processing. Our results showed moderate to strong relationships exist between somatosensory measures and (1) the response to visual perturbation for CP/TD, and (2) the improvement in the response with SR intervention for CP, regardless of More/Less Affected Side. Thus, impairments in joint position sense, vibration and two-point discrimination could all predict the response to visual perturbations, as well as the improvement in the response from SR stimulation for CP.

These results support our hypothesis and are not surprising. Although the sensory tests measure different “senses” such as vibration and joint position sense, the sensory receptors involved in perceiving these sensations are overlapping. For example, vibrotactile stimulation activates the Pacinian corpuscles, which are the primary mechanoreceptors stimulated by cutaneous vibration sensation, as well as the ankle joint proprioceptors (Mildren & Bent, 2016), indicating a complex interplay between sensory stimuli and sensory receptors. Further, the fact that sensory tests show strong correlation with the response to visual perturbations over all CP/TD group and More/Less Affected Side combinations and with the improvement in the response to SR for CP may indicate a largely central cause for the observed sensory deficits in individuals with CP, such as aberrant sensory processing and integration of sensory input by the central nervous system, rather than deficits in peripheral nervous system involving the sensory receptors. Improvement in the response to visual perturbations with a peripheral stimulation at the lower extremities, however, does point towards some potential involvement of the peripheral nervous system, e.g., deficits in perceiving sensory input. While the sensory deficits in individuals with CP are primarily due to an injury of the central nervous system, limited environmental exploration and learning experiences may secondarily lead to impairments in the somatosensory system (Hoon et al., 2009; Kurz et al., 2014). Thus, while the peripheral stimulation using a sensory-centric therapy such as SR does not directly address the primary brain injury, it addresses the secondary somatosensory deficits arising from reduced environmental exploration during development.

Our finding that ankle joint proprioception is strongly correlated with response to visual perturbation is consistent with prior research on standing postural sway in children with CP, which showed that ankle joint position sense was strongly correlated with standing balance measured via the BESTest (Zarkou et al., 2020). Furthermore, our result show that greater deficits in vibration sense correlated strongly with improvement in the response to visual perturbation after receiving SR stimulation. This is particularly interesting in light of prior work that showed that vibration sensation at the first metatarsal head showed a significantly strong correlation with motor performance and spatiotemporal gait parameters (Zarkou et al., 2020). Thus, these findings underscore the role of vibration sensation and ankle joint position sense in maintaining upright balance during walking and imply that the greater the somatosensory deficits, the worse the balance responses to challenges during walking and, in turn, the higher the potential for improvement with sensory-centric interventions.

Our study has some limitations that should be considered while interpreting the results. First, this study is a secondary analysis of data collected for two different experimental protocols. The sample size was selected based on the power analysis for the primary analyses designed to test between group differences between CP and TD in the responses to visual perturbations and in improvements to SR stimulation and was not selected to sufficiently power the regression analyses. Second, our study only included individuals with GMFCS levels I and II. Thus, individuals with higher GMFCS levels may have a different relationship between somatosensory deficits and visual reliance.

Collectively, this study provides preliminary evidence that sensory deficits play a role in walking balance problems in individuals with CP and the potential for improvement with sensory interventions. Our evidence is of particular clinical relevance because current interventions provide limited to mixed results(Blumetti et al., 2012; Pin et al., 2006; Taylor et al., 2013). Our results show that performing a simple battery of sensory tests to evaluate joint position sense, vibration and two-point discrimination could identify individuals who are most likely to be heavily reliant on vision for balance control. Somatosensory tests can be used to identify individuals who are most likely to respond favorably to sensory-centric interventions. Overall, our research supports the inclusion of sensory based interventions in addition to the current motor-centric treatments, thus providing a more comprehensive and holistic approach to balance rehabilitation in individuals with CP.

## Data Availability

All data produced in the present study are available upon reasonable request to the authors

## 5. ACKNOWLEDGEMENTS

AS was supported in part by the Foundation for Physical Therapy Research Promotion of Doctoral Studies II, American Society of Biomechanics Grant-in-Aid, and the University of Delaware Graduate College through the Unidel Distinguished Graduate Scholar Award. HR was supported by the Parkinson’s Foundation (PF-JFA-2036) and the National Science Foundation (NSF CRCNS 1822568). The authors have stated that theyhad no interests which may be perceived as posing a conflict or bias.

## REFERENCES

Barela, J. A., Focks, G. M. J., Hilgeholt, T., Barela, A. M. F., de P. Carvalho, R., & Savelsbergh, G. J. P. (2011). Perception-action and adaptation in postural control of children and adolescents with cerebral palsy. Research in Developmental Disabilities, 32(6), 2075–2083. 10.1016/j.ridd.2011.08.018

Bell-Krotoski, J., Weinstein, S., & Weinstein, C. (1993). Testing Sensibility, Including Touch-Pressure, Two-point Discrimination, Point Localization, and Vibration. Journal of Hand Therapy, 6(2), 114–123. 10.1016/S0894-1130(12)80292-4

Blumetti, F. C., Wu, J. C. N., Bau, K. V., Martin, B., Hobson, S. A., Axt, M. W., & Selber, P. (2012). Orthopedic surgery and mobility goals for children with cerebral palsy GMFCS level IV: what are we setting out to achieve? Journal of Children’s Orthopaedics, 6(6), 485–490. 10.1007/s11832-012-0454-7

Boyer, E. R., & Patterson, A. (2018). Gait pathology subtypes are not associated with self-reported fall frequency in children with cerebral palsy. Gait & Posture, 63, 189–194. 10.1016/j.gaitpost.2018.05.004

Cordo, P., Inglis, J. T., Verschueren, S., Collins, J. J., Merfeld, D. M., Rosenblum, S., Buckley, S., & Moss, F. (1996). Noise in human muscle spindles. Nature, 383(6603), 769–770. 10.1038/383769a0

Damiano, D. L., Wingert, J. R., Stanley, C. J., & Curatalo, L. (2013). Contribution of hip joint proprioception to static and dynamic balance in cerebral palsy: a case control study. Journal of Neuroengineering and Rehabilitation, 10(1), 57. 10.1186/1743-0003-10-57

Franz, J. R., Francis, C. A., Allen, M. S., O’Connor, S. M., & Thelen, D. G. (2015). Advanced age brings a greater reliance on visual feedback to maintain balance during walking. Human Movement Science, 40, 381–392. 10.1016/j.humov.2015.01.012

Gravelle, D. C., Laughton, C. A., Dhruv, N. T., Katdare, K. D., Niemi, J. B., Lipsitz, L. A., & Collins, J. J. (2002). Noise-enhanced balance control in older adults. Neuroreport, 13(15), 1853–1856. 10.1097/00001756-200210280-00004

Hoon, A. H., Stashinko, E. E., Nagae, L. M., Lin, D. D., Keller, J., Bastian, A., Campbell, M. L., Levey, E., Mori, S., & Johnston, M. V. (2009). Sensory and motor deficits in children with cerebral palsy born preterm correlate with diffusion tensor imaging abnormalities in thalamocortical pathways. Developmental Medicine and Child Neurology, 51(9), 697–704. 10.1111/j.1469-8749.2009.03306.x [doi]

Jeffries, L., Fiss, A., McCoy, S. W., & Bartlett, D. J. (2016). Description of Primary and Secondary Impairments in Young Children With Cerebral Palsy. Pediatric Physical Therapy, 28(1), 7. 10.1097/PEP.0000000000000221

Kurz, M. J., Heinrichs-Graham, E., Arpin, D. J., Becker, K. M., & Wilson, T. W. (2014). Aberrant synchrony in the somatosensory cortices predicts motor performance errors in children with cerebral palsy. Journal of Neurophysiology, 111(3), 573–579. 10.1152/jn.00553.2013 [doi]

Kurz, M. J., Heinrichs-Graham, E., Becker, K. M., & Wilson, T. W. (2015). The magnitude of the somatosensory cortical activity is related to the mobility and strength impairments seen in children with cerebral palsy. Journal of Neurophysiology, 113(9), 3143–3150. 10.1152/jn.00602.2014

Mildren, R. L., & Bent, L. R. (2016). Vibrotactile stimulation of fast-adapting cutaneous afferents from the foot modulates proprioception at the ankle joint. Journal of Applied Physiology, 120(8), 855–864. 10.1152/japplphysiol.00810.2015

Moss, F., Ward, L. M., & Sannita, W. G. (2004). Stochastic resonance and sensory information processing: a tutorial and review of application. Clinical Neurophysiology: Official Journal of the International Federation of Clinical Neurophysiology, 115(2), 267–281.

Pin, T., Dyke, P., & Chan, M. (2006). The effectiveness of passive stretching in children with cerebral palsy. Developmental Medicine and Child Neurology, 48(10), 855–862. 10.1017/S0012162206001836

Priplata, A. A., Patritti, B. L., Niemi, J. B., Hughes, R., Gravelle, D. C., Lipsitz, L. A., Veves, A., Stein, J., Bonato, P., & Collins, J. J. (2006). Noise-enhanced balance control in patients with diabetes and patients with stroke. Annals of Neurology, 59(1), 4–12. 10.1002/ana.20670

Ribot-Ciscar, E., Hospod, V., & Aimonetti, J.-M. (2013). Noise-enhanced kinaesthesia: a psychophysical and microneurographic study. Experimental Brain Research, 228(4), 503–511. 10.1007/s00221-013-3581-6

Ross, S. E., & Guskiewicz, K. M. (2006). Effect of Coordination Training With and Without Stochastic Resonance Stimulation on Dynamic Postural Stability of Subjects With Functional Ankle Instability and Subjects With Stable Ankles. Clinical Journal of Sport Medicine, 16(4). https://journals.lww.com/cjsportsmed/Fulltext/2006/07000/Effect_of_Coordination_Training_With_and_Without.7.aspx

Ross, S. E., Linens, S. W., Wright, C. J., & Arnold, B. L. (2013). Customized Noise-Stimulation Intensity for Bipedal Stability and Unipedal Balance Deficits Associated With Functional Ankle Instability. Journal of Athletic Training, 48(4), 463–470. 10.4085/1062-6050-48.3.12

Sansare, A., Arcodia, M., Lee, S. C. K., Jeka, J., & Reimann, H. (2022). Individuals with cerebral palsy show altered responses to visual perturbations during walking. Frontiers in Human Neuroscience, 16. 10.3389/fnhum.2022.977032

Sansare, A., Reimann, H., Crenshaw, J., Arcodia, M., Verma, K., & Lee, S. C. K. (2023). Subthreshold electrical noise alters walking balance control in individuals with cerebral palsy. Gait & Posture, 106, 47–52. 10.1016/j.gaitpost.2023.08.008

Sansare Ashwini, Arcodia Maelyn, Lee Samuel, Jeka John, & Reimann Hendrik. (n.d.). Immediate application of low-intensity electrical noise reduced responses to visual perturbations during walking in individuals with cerebral palsy. Journal of NeuroEngineering and Rehabilitation. 10.21203/rs.3.rs-2824563/v1

Stephen, D. G., Wilcox, B. J., Niemi, J. B., Franz, J., Casey Kerrigan, D., & D’Andrea, S. E. (2012). Baseline-dependent effect of noise-enhanced insoles on gait variability in healthy elderly walkers. Gait & Posture, 36(3), 537–540. 10.1016/J.GAITPOST.2012.05.014

Taylor, N. F., Dodd, K. J., Baker, R. J., Willoughby, K., Thomason, P., & Graham, H. K. (2013). Progressive resistance training and mobility-related function in young people with cerebral palsy: a randomized controlled trial. Developmental Medicine and Child Neurology, 55(9), 806–812. 10.1111/dmcn.12190

Temlett, J. A. (2009). An assessment of vibration threshold using a biothesiometer compared to a C128-Hz tuning fork. Journal of Clinical Neuroscience, 16(11), 1435–1438. 10.1016/J.JOCN.2009.03.010

Wingert, J. R., Burton, H., Sinclair, R. J., Brunstrom, J. E., & Damiano, D. L. (2009). Joint-position sense and kinesthesia in cerebral palsy. Archives of Physical Medicine and Rehabilitation, 90(3), 447–453. 10.1016/j.apmr.2008.08.217

Yu, Y., Lauer, R. T., Tucker, C. A., Thompson, E. D., & Keshner, E. A. (2018). Visual dependence affects postural sway responses to continuous visual field motion in individuals with cerebral palsy. Developmental Neurorehabilitation, 21(8), 531–541. 10.1080/17518423.2018.1424265

Zarkou, A., Lee, S. C. K., Prosser, L. A., & Jeka, J. J. (2020). Foot and Ankle Somatosensory Deficits Affect Balance and Motor Function in Children With Cerebral Palsy. Frontiers in Human Neuroscience, 14. 10.3389/fnhum.2020.00045

